# Performance of microbiological tests for tuberculosis diagnostic according to the type of respiratory specimen: a 10-year retrospective study

**DOI:** 10.1101/2022.12.27.22283924

**Authors:** Marc-Olivier Boldi, Justin Denis-Lessard, Rina Neziri, René Brouillet, Christophe von-Garnier, Valérie Chavez, Jesica Mazza-Stalder, Katia Jaton, Gilbert Greub, Onya Opota

**Affiliations:** Faculty of Business and Economics, University of Lausanne, Switzerland; Institute of Microbiology, Lausanne University and University Hospital of Lausanne, Lausanne, Switzerland; Pulmonary Division, Lausanne University Hospital, University of Lausanne, Lausanne, Switzerland; Infectious Diseases Service, Lausanne University and University Hospital of Lausanne, Lausanne, Switzerland

**Keywords:** Tuberculosis, microscopy, PCR, mycobacterial culture, bronchoalveolar lavage, bronchial aspirate, sputum, induced-sputum, acid-fast bacilli (AFB)

## Abstract

**Background:** The microbial diagnosis of tuberculosis (TB) remains challenging and relies on multiple microbiological tests performed on different clinical specimens. Polymerase chain reactions (PCRs), introduced in the last decades has had a significant impact on the diagnosis of TB. However, questions remain about the use of PCRs in combination with conventional tests for TB, namely microscopy and culture. We aimed to determine the performance of microscopy, culture and PCR for the diagnosis of pulmonary tuberculosis according to the type of clinical specimen in order to improve the diagnostic yield and to avoid unnecessary, time and labor-intensive tests.

**Methods:** We conducted a retrospective study (2008-2018) on analysis (34’429 specimens, 14’358 patients) performed in our diagnostic laboratory located in the Lausanne University Hospital to compare the performance of microbiological tests on sputum, induced sputum, bronchial aspirate and bronchoalveolar lavage (BAL). We analysed the performance using a classical “per specimen” approach and a “per patient” approach for paired specimens collected from the same patient.

**Results:** The overall sensitivities of microscopy, PCR and culture were 0.523 (0.489, 0.557), 0.798 (0.755, 0.836) and 0.988 (0.978, 0.994) and the specificity were 0.994 (0.993, 0.995), 1 (0.999, 1) and 1 (1, 1). Microscopy displayed no significant differences in sensitivity according to the type of sample. The sensitivities of PCR for sputum, induced sputum, bronchial aspirate and BAL were, 0.821 (0.762, 0.871), 0.643 (0.480, 0.784), 0.837 (0.748, 0.904) and 0.759 (0.624, 0.865) respectively and the sensitivity of culture were, 0.993 (0.981, 0.998), 0.980 (0.931, 0.998), 0.965 (0.919, 0.988), and 1 (0.961, 1) respectively. Pairwise comparison of specimens collected from the same patient reported a significantly higher sensitivity of PCR on bronchial aspirate over BAL (p < 0.001) and sputum (p < 0.05) and a significantly higher sensitivity of culture on bronchial aspirate over BAL (p < 0.0001).

**Conclusions:** PCR displayed a higher sensitivity and specificity than microscopy for all respiratory specimens, a rational for a smear-independent PCR-based approach to initiate tuberculosis microbial diagnostic. The diagnosis yield of bronchial aspirate was higher than BAL. Therefore, PCR should be systematically performed also on bronchial aspirates when available.

## 1. Introduction

With over ten million new cases in 2020 and about 1,5 million deaths, tuberculosis represents a major public health concern [1]. Rapid and reliable diagnosis is important to reduce morbidity and mortality associated with tuberculosis and to control transmission. When tuberculosis is suspected based on clinical symptoms, epidemiological information and radiological findings, microbial confirmation is key to establish the diagnosis. Despite progress during the last decades, the microbiological diagnosis of tuberculosis continues to be a challenge particularly in paucibacillary disease. Historically, the diagnosis of tuberculosis was based on microscopy and culture. Mycobacterial culture represents the reference method due to a low limit of detection (< 10 organisms for liquid cultures) and because it gives access to the strain for phenotypic antibiotic susceptibility test [1,2]. However, culture is challenging due to the slow growth of *M. tuberculosis* and because it requires biosafety level three (BSL3) laboratories [3,4]. Microscopy based on the visualization of acid-fast bacilli provides rapid results (<30 minutes) but has a limited sensitivity and specificity (limit of detection between 10^3^ and 10^4^ bacilli per ml) [5]. In order to increase their sensitivity, these tests may need to be repeated over several clinical specimen [1,6,7].

More recently, molecular diagnosis, in particular polymerase chain reaction (PCR) have improved the diagnosis of tuberculosis with a limit of detection between 10 and 10^3^ colony forming units per ml and a turnaround time between two to six hours [2,6,8,9]. PCR was initially available in laboratories specialized in molecular diagnostics, through methods developed in-house assays [10,11]. Commercial all-inclusive systems, such as the GeneXpert system, now allow a greater number of laboratories to perform this analysis independently of a specialized infrastructure [6,8,9,12]. The GeneXpert system not only improved the initial diagnostic of tuberculosis but can be used to assess patient’s infectious potential, on the basis of the semi-quantitative results [2,5,8]. In addition, rapid molecular test also shortens airborne isolation for hospitalized patients with presumptive tuberculosis [13]

More than 70% of the tuberculosis infections are pulmonary tuberculosis, for which sputum is the usual specimen collected in adults and older children who are able to collaborate. Other respiratory specimens can be considered when patients are not able to provide sputum or to increase microbial diagnostic yield [10]. This includes induced sputum, obtained by nebulization of sterile hypertonic saline (3% or 7% saline solution inhaled) followed by coughing and expectoration of airway secretions, bronchial aspirate, and bronchoalveolar lavages [14,15].

In this study, we aimed to assess the performance of the different tests for the microbial diagnostic of tuberculosis according to the type of clinical specimen. Providing robust updated data may enable to choose optimal combination of test and specimen to: i) reach the maximum sensitivity, specificity and negative and predicative value, ii) prioritize the tests and specimens in the situation of limited resources or shortage of material and iii) reduce unnecessary costs.

There is no standard method to address the performance of diagnostic tests, particularly for tuberculosis where several microbiological tests on multiple clinical samples are frequently required. We applied data analytics methods that integrate multiple parameters, including, the type of microbiological test, the type of specimens, the sampling period, and the patients. In this study, we performed both a classical “per specimen” method to determine the performance of each diagnostic test and a “per patient” approach comparing paired specimens collected from the same patient.

This study provides data to establish diagnostic stewardship guidelines and diagnostic protocols. These data will help to establish more effective strategies to diagnose pulmonary tuberculosis in order to increase the rate of documentation, to accelerate the diagnosis and avoid unnecessary testing. In addition, it should provide analytical strategies that may also be suitable to study other infectious diseases while keeping associated medical, social and economic costs to a strict minimum.

## 2. Material and Methods

### 2.1 Study Design and data

Our laboratory is located in the Lausanne University Hospital (CHUV), a 1’500 beds tertiary-care hospital in a low-tuberculosis-prevalence country (Switzerland), with approximately six new cases per year per 100,000 population (Federal Office of Public Health; http://www.bag.admin.ch/). The data included microbiology analyses for patients with suspected mycobacterial infection from 2008 to 2018. They were automatically extracted from the Laboratory Information System (MOLIS, CGM).

For all specimens, information regarding the microbial diagnostic of mycobacteria were extracted. This included information regarding microscopy, PCR, cultures, molecular and phenotypic resistance genes together with the type of specimen and the date of collection. Each specimen was given a unique coded number. Similarly, each patient was given a unique coded number. The database was generated to allow analysis by date of sampling and by patients (see Section 2.4). For the microbial diagnostic of the disease, we generated a composite gold standard including microbiological findings and epidemiological and clinical data (see Section 2.3). The final database included 34’429 specimens corresponding to 14’358 patients, including 8’587 sputum, 2’257 induced sputum, 8’610 bronchial aspirate and 4’576 BAL.

### 2.2 Clinical specimens and mycobacterial diagnostic test

This study focuses on three microbial diagnostic tests commonly used for the diagnostic of mycobacterial infections, namely (i) microscopy (also named smear microscopy) for the direct detection of acid-fast bacilli (AFB) in clinical specimen, (ii) PCR for the detection of DNA of *M. tuberculosis* complex directly from clinical specimen and (iii) mycobacterial culture. Microscopy consists in acid-fast bacillus staining achieved through a fluorescent auramine-thiazine red staining on a heat-fixed smear as described in [8]. PCR consisted of either an in-house TaqMan PCR targeting the multicopy *M. tuberculosis* IS6110 sequence, named PCR “MYTU” [11] or using the all-inclusive rapid molecular test Xpert MTB/RIF and Xpert MTB/RIF Ultra (herafter named Xpert) (Cepheid, Ca, USA) [6,8,16]. Mycobacterial culture was achieved using mycobacterial growth indicator tube (MGIT) that consists in culture tubes containing tris 4, 7-diphenyl-1, 10-phenonthroline ruthenium chloride pentahydrate, an oxygen-quenched fluorochrome embedded in silicone at their bottom. Utilization of free oxygen for growing bacteria alleviates the fluorochrome quenching, resulting in fluorescence within the tube that can be visualized under UV light, as explained in a previous publication [8].

All the microbial analyses were performed on the same sample after splitting it for AFB staining, Xpert analysis and mycobacterial culture [8]. Sputum, induced sputum, bronchial aspirate and bronchoalveolar lavage were processed as previously described [8]. In particular, samples with a volume exceeding 3 mL were concentrated by centrifugation (30 minutes, 3000 *g*). In addition, to increase the homogeneity of the sample before smear preparation, purulent sputum or bronchial aspirates were solubilized with the mucolytic agent N-acetyl-L-cysteine (2% v/v pH 6.8) [8].

### 2.3 Composite gold standard

Mycobacterial culture is the gold standard for the diagnosis of *M. tuberculosis* because of its lowest limit of detection (LOD < 10 organisms). However, it can be impaired by situation that affect mycobacterial growth such as the introduction of an antibiotic treatment before sampling [2]. We therefore used a composite gold standard based on microbiological results and clinical data. Discrepant results in the diagnostic of active tuberculosis, especially specimen with positive MTBC PCR and negative culture for *M. tuberculosis* complex, were manually cured based on clinical and epidemiological data found in medical records. Specimens for which the culture was contaminated by bacteria of the flora were excluded. Specimens with culture positive with nontuberculous mycobacteria qualified as “MOTT” (Mycobacteria other than tuberculosis) were considered negative for *M. tuberculosis*. This resulted in the “Gold Standard” (GS) reference.

### 2.4 Performance of the test depending on the clinical specimens

We used two different approaches to determine the performance of microbiological tests depending on the clinical specimens. We first used a common “per specimen” approach to determine the global performance of microscopy, PCR, and culture according to the four types of specimens and independently of the patient using the GS as reference. Then, in order to provide more robust data we performed a “per patient” approach. It consists in pairwise comparison for specimens collected the same day or during a window of 72 hours for the same patient; indeed, samples from the same patient might not be collected the same day. For each patient, the first sample was paired to the following sample if it was of a different type and within a 72-hour window. All the combinations of the four types of specimens were analyzed using this pairwise approach i) to measure the dependence between each pair of types of specimens and ii) to calculate the performance of one sample type to predict a positive of any of the two sample types.

### 2.5 Statistics

The performance measures consist in accuracy, sensitivity, specificity, positive and negative predictive values. They were computed using GS as reference. Their respective 95% confidence intervals were computed using the Clopper–Pearson method. The comparisons of proportions were assessed with a two-sided proportion test with Yates’ continuity correction. The dependence between pairs of types of specimens for the “per patient” approach is measured by the Cohen’s Kappa.

### 2.6 Ethics committee approval

This study was approved by the relevant ethics committee, the *Commission Cantonale d’Éthique de la Recherche sur l’Être Humain* (*CER-2020-00136)*.

## 3. Results

### 3.1 Global performance of microscopy, PCR and culture for the diagnostic of pulmonary tuberculosis

We first determined the global performance of microscopy. The sensitivity, 0.523 (0.489 – 0.557) and the PPV, 0.767 (0.730, 0.800) of microscopy were limited. The specificity, 0.994 (0.993, 0.995), and the NPV 0.982 (0.981, 0.984) were high but must be interpreted according to the low prevalence of microscopy positive specimen 0.036 (0.034, 0.038) (Table 1 and Table S1).

**Table 1.** Performance of smear microscopy for the diagnostic of pulmonary tuberculosis according of the type of specimen.

|  |  |  |  |  |  |  |
| --- | --- | --- | --- | --- | --- | --- |
|  | Accuracy | Sensitivity | Specificity | Prevalence | PPV | NPV |
| **All specimens** | 0.977<br>(0.975, 0.979)<br>23219/23763 | 0.523<br>(0.489, 0.557)<br>447/855 | 0.994<br>(0.993, 0.995)<br>22772/22908 | 0.036<br>(0.034, 0.038)<br>855/23763 | 0.767<br>(0.730, 0.800)<br>447/583 | 0.982<br>(0.981, 0.984)<br>22772/23180 |
| **Sputum** | 0.967<br>(0.963, 0.971)<br>8132/8409 | 0.605<br>(0.562, 0.647)<br>320/529 | 0.991<br>(0.989, 0.993)<br>7812/7880 | 0.063<br>(0.058, 0.068)<br>529/8409 | 0.825<br>(0.783, 0.861)<br>320/388 | 0.974<br>(0.970, 0.977)<br>7812/8021 |
| **Induced sputum** | 0.966<br>(0.957, 0.973)<br>2119/2194 | 0.362<br>(0.265, 0.467)<br>34/94 | 0.993<br>(0.988, 0.996)<br>2085/2100 | 0.043<br>(0.035, 0.052)<br>94/2194 | 0.694<br>(0.546, 0.817)<br>34/49 | 0.972<br>(0.964, 0.979)<br>2085/2194 |
| **Bronchial aspirate** | 0.985<br>(0.982, 0.988)<br>8463/8591 | 0.362<br>(0.283, 0.447)<br>51/141 | 0.996<br>(0.994, 0.997)<br>8412/8450 | 0.016<br>(0.014, 0.019)<br>141/8591 | 0.573<br>(0.464, 0.677)<br>51/89 | 0.989<br>(0.987, 0.991)<br>8412/8502 |
| **BAL** | 0.986<br>(0.982, 0.989)<br>4505/4569 | 0.462<br>(0.356, 0.569)<br>42/91 | 0.997<br>(0.994, 0.998)<br>4463/4478 | 0.020<br>(0.016, 0.024)<br>91/4569 | 0.737<br>(0.603, 0.845)<br>42/57 | 0.989<br>(0.986, 0.992)<br>4463/4512 |
| All p.values in supplementary material. |  |  |  |  |  |  |

Regarding PCR, we first estimated the individual performance of the in-house TaqMan PCR and the rapid molecular test Xpert. The sensitivity of the in-house TaqMan PCR, 0.799 (0.743, 0.848) and Xpert 0.812 (0.760, 0.858) were not significantly different (Table S2 and Table S3). The specificity for both the in-house TaqMan PCR >0.999 (0.999, 1) and Xpert 0.999 (0.997, 1) were both very high. The NPV were also high but probably increased by the low prevalence of PCR positive specimen (Table S7). Because the two PCR tests displayed similar performance, we considered them as equal for the rest of the study and grouped them as “PCR”. The global performance of PCR were: sensitivity 0.798 (0.755, 0.836), specificity 1 (0.999, 1), PPV 0.997 (0.983, 1) and NPV 0.988 (0.985, 0.990) (Table 2 and S2).

**Table 2.**
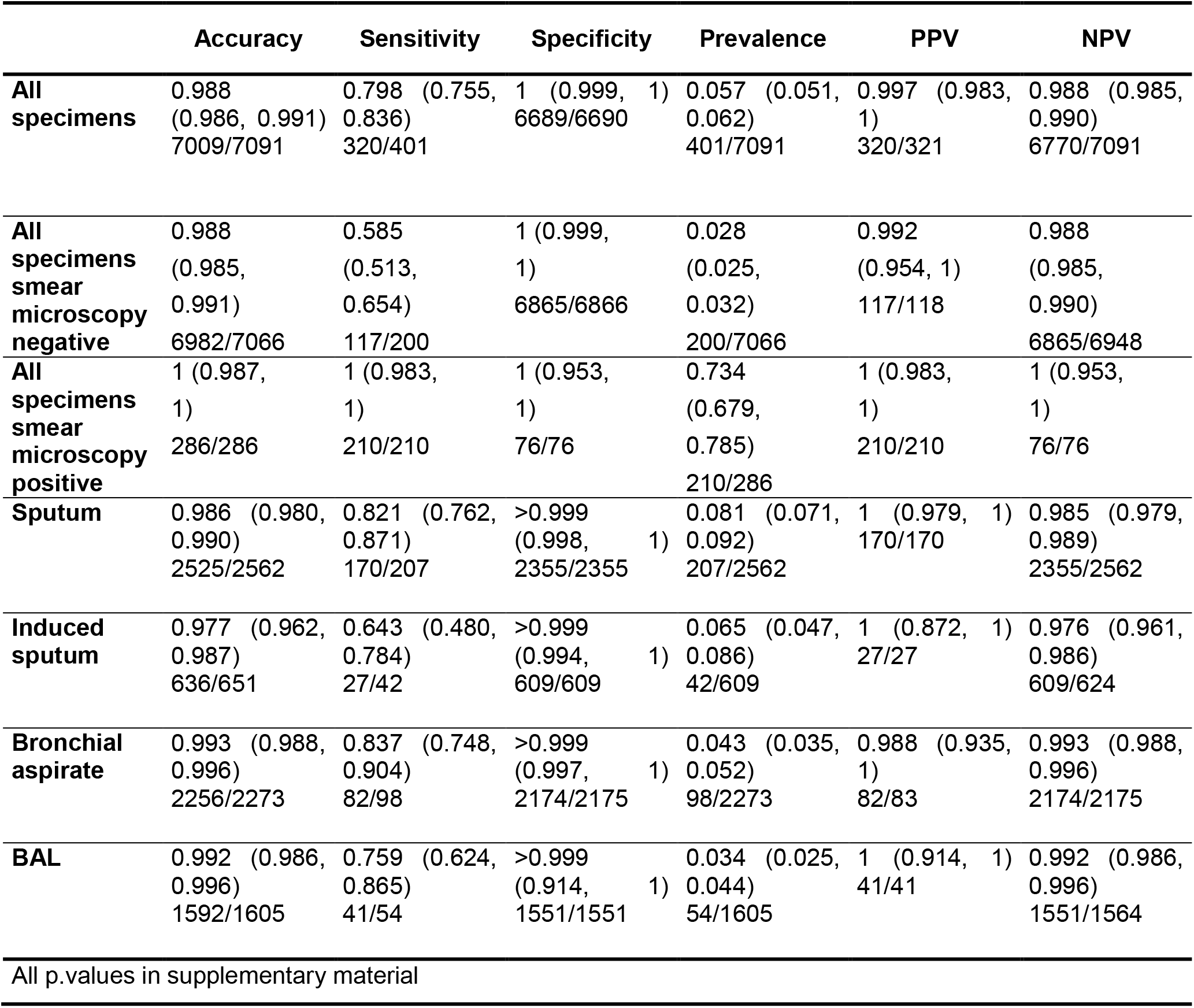
Performance of PCR for the diagnostic of pulmonary tuberculosis according to the type of specimen.

The culture displayed the highest performance for the diagnostic of tuberculosis using the GS reference with a sensitivity of 0.988 (0.978, 0.994) and a specificity of 1 (1, 1) (Table 3 and Table S3). In summary when considering all the clinical specimen, microscopy displays limited sensitivity, PCR displays a higher sensitivity than microscopy and an excellent specificity and culture displayed the highest sensitivity and specificity.

**Table 3:** Performance of culture for the diagnostic of pulmonary tuberculosis according to the type of specimen.

|  |  |  |  |  |  |  |
| --- | --- | --- | --- | --- | --- | --- |
|  | Accuracy | Sensitivity | Specificity | Prevalence | PPV | NPV |
| All specimens | >0.999<br>(0.999, 1)<br>24019/24030 | 0.988<br>(0.978, 0.994)<br>871/882 | 1 (1, 1)<br>23148/23148 | 0.037<br>(0.034, 0.039)<br>882/24030 | 1 (0.996, 1)<br>871/871 | 1 (0.999, 1)<br>23148/23159 |
| Sputum | >0.999<br>(0.999, 1)<br>8583/8587 | 0.993<br>(0.981, 0.998)<br>543/547 | 1 (1, 1)<br>8040/8040 | 0.064<br>(0.059, 0.069)<br>547/8587 | 1 (0.993, 1)<br>543/543 | 1 (0.999, 1)<br>8040/8044 |
| Induced sputum | 1 (0.997, 1)<br>2255/2257 | 0.980<br>(0.931, 0.998)<br>100/102 | 1 (0.998, 1)<br>2155/2155 | 0.045<br>(0.037, 0.055)<br>102/2155 | 1 (0.964, 1)<br>100/100 | 1 (0.997, 1)<br>2155/2157 |
| Bronchial aspirate | >0.999<br>(0.999, 1)<br>8605/8610 | 0.965<br>(0.919, 0.988)<br>136/141 | 1 (1, 1)<br>8469/8469 | 0.016<br>(0.014, 0.019)<br>141/8610 | 1 (0.973, 1)<br>136/136 | 0.999 (0.999, 1)<br>8469/8474 |
| BAL | 1 (0.999, 1)<br>4576/4576 | 1 (0.961, 1)<br>92/92 | 1 (0.999, 1)<br>4484/4484 | 0.020<br>(0.016, 0.025)<br>92/4576 | 1 (0.961, 1)<br>92/92 | 1 (0.999, 1)<br>4484/4484 |
| All p.values in supplementary material |  |  |  |  |  |  |

### 3.3 Performance of microbiological tests according to the type of specimen using a “per sample” approach or using “per patient” pairwise comparisons

We next measured the dependence between the specimens using the “per patient” approach and the Cohen’s kappa. Though the Cohen’s kappa is itself difficult to interpret and the confidence intervals shown in the table were not corrected for multiple comparisons, a large value of kappa means that the two sample bring the same information to some extent and thus provides little “complementary” information and are so-called “supllementary”, i.e not really needed. Conversely, a kappa close to zero means that the two sample are “complementary”. The results are shown in Table S8, S9, S10 and S11. The data suggested that for each technique, microscopy, PCR, and culture, most samples are supplementary (i.e. not all needed), except i) for microscopy, induced sputum versus bronchial aspirate and BAL, ii) for PCR, sputum versus induced sputum and iii) for the culture, induced sputum versus BAL. Because of their low robustness, we do not want to over interpret these results by concluding that the so-called “supplementary” tests don’t need to be performed, but this question has to be tackled in additional work since it is however to our knowledge the first study showing such high dependence & redundancy of the various tests.

### 3.4 Performance of microscopy according to the type of respiratory specimen

Using the classical “per specimen” approach the sensitivities of microscopy for all specimen, sputum, induced sputum, bronchial aspirate and BAL were 0.523 (0.489, 0.557), 0.605 (0.562, 0.647), 0.362 (0.265, 0.467), 0.362 (0.283, 0.447) and 0.462 (0.356, 0.569) respectively. The sensitivity of sputum was higher than induced sputum (p-value < 0.0001) and bronchial aspirate (p-value < 0.0001) (Table 4 and S4). However, using the “per patient” comparison approach, for paired specimens collected within a window of 72h for the same patient, no significant difference was observed for the various specimens (Table 4,S4 and S9). Altogether, these data suggested a limited benefit of microscopy for tuberculosis microbial diagnosis in a low prevalence setting.

**Table 4.** Sensitivity of microscopy to predict tuberculosis according to the type of specimen using a 72-hours pairing window in the same patient.

|  |  |  |
| --- | --- | --- |
|  | Sensitivity | NPV |
| **Sputum versus induced sputum** |  |  |
| **Sputum** | 0.923 (0.640, 0.998) 12/13 <sup>n.s</sup> | 0.997 (0.986, 1) 390/391 <sup>n.s</sup> |
| **Induced Sputum** | 0.769 (0.462, 0.950) 10/13 <sup>n.s</sup> | 0.992 (0.978, 0.998) 390/393 <sup>n.s</sup> |
| **Sputum versus bronchial aspirate** |  |  |
| **Sputum** | 0.692 (0.386, 0.909) 9/13 <sup>n.s</sup> | 0.988 (0.970, 0.997) 337/341 <sup>n.s</sup> |
| **Bronchial aspirate** | 0.846 (0.546, 0.981) 11/13 <sup>n.s</sup> | 0.994 (0.979, 0.999) 337/339 <sup>n.s</sup> |
| **Sputum versus BAL** |  |  |
| **Sputum** | 1 (0.631, 1) 8/8 <sup>n.s</sup> | 1 (0.987, 1) 280/280 <sup>n.s</sup> |
| **BAL** | 0.750 (0.349, 0.968) 6/8 <sup>n.s</sup> | 0.993 (0.975, 0.999) 280/282 <sup>n.s</sup> |
| **Induced sputum versus bronchial aspirate** |  |  |
| **Induced Sputum** | 0.500 (0.013, 0.987) 1/2 <sup>n.s</sup> | 0.982 (0.904, 1) 55/56 <sup>n.s</sup> |
| **Bronchial aspirate** | 0.500 (0.013, 0.987) 1/2 <sup>n.s</sup> | 0.982 (0.904, 1) 55/56 <sup>n.s</sup> |
| **Induced sputum versus BAL** |  |  |
| **Induced Sputum** | 0.333 (0.008, 0.906) 1/3 <sup>n.s</sup> | 0.962 (0.868, 0.995) 50/52 <sup>n.s</sup> |
| **BAL** | 0.667 (0.094, 0.992) 2/3 <sup>n.s</sup> | 0.980 (0.896, 1) 50/51 <sup>n.s</sup> |
| **Bronchial aspirate versus BAL** |  |  |
| **Bronchial aspirate** | 0.881 (0.744, 0.960) 37/42 <sup>n.s</sup> | 0.999 (0.997, 1.000) 3528/3533 <sup>n.s</sup> |
| **BAL** | 0.714 (0.554, 0.843) 30/42 <sup>n.s</sup> | 0.997 (0.994, 0.998) 3528/3540 <sup>n.s</sup> |
| All p.values in supplementary material |  |  |

### 3.5 Performance of PCR according to the type of respiratory specimen

Using the classical “per specimen” approach the sensitivities of PCR for all specimen, sputum, induced sputum, bronchial aspirate and BAL were 0.798 (0.755, 0.836), 0.821 (0.762, 0.871), 0.643 (0.480, 0.784), 0.837 (0.748, 0.904) and 0.759 (0.624, 0.865) respectively. Using the “per patient” pairwise comparison approach, no significant difference in sensitivity was seen between sputum and induced sputum. Using this approach, we found that the sensitivity of bronchial aspirate, was significantly higher than BAL with respectively 0.974 (0.865, 0.999) and 0.564 (0.396, 0.722) (p. value 0.0003). The sensitivity of bronchial aspirate was also higher than sputum with respectively 1 (0.753, 1) *versus* 0.385 (0.139, 0.684) (p-value = 0.017) (Table 5,S5 and S10).

**Table 5.** Sensitivity of PCR to predict tuberculosis according to the type of specimen using a 72-hours pairing window in the same patient.

|  |  |  |
| --- | --- | --- |
|  | **Sensitivity** | **NPV** |
| **Sputum versus induced sputum** |  |  |
| **Sputum** | 0.667 (0.094, 0.992) 2/3 <sup>n.s</sup> | 0.992 (0.958, 1) 129/130 <sup>n.s</sup> |
| **Induced sputum** | 0.667 (0.094, 0.992) 2/3 <sup>n.s</sup> | 0.992 (0.958, 1) 129/130 <sup>n.s</sup> |
| **Sputum versus bronchial aspirate** |  |  |
| **Sputum** | 0.385 (0.139, 0.684) 5/13 * | 0.953 (0.909, 0.979) 161/169 <sup>n.s</sup> |
| **Bronchial aspirate** | 1 (0.753, 1) 13/13 * | 1 (0.977, 1) 161/161 <sup>n.s</sup> |
| **Sputum versus BAL** |  |  |
| **Sputum** | 0.600 (0.147, 0.947) 3/5 <sup>n.s</sup> | 0.985 (0.947, 0.998) 132/134 <sup>n.s</sup> |
| **BAL** | 1 (0.478, 1) 5/5 <sup>n.s</sup> | 1 (0.478, 1) 132/132 <sup>n.s</sup> |
| **Induced sputum versus bronchial aspirate** |  |  |
| **Induced sputum** | 0.857 (0.421, 0.996) 6/7 <sup>n.s</sup> | 0.968 (0.833, 0.999) 30/31 <sup>n.s</sup> |
| **Bronchial aspirate** | 0.714 (0.290, 0.963) 5/7 <sup>n.s</sup> | 0.938 (0.792, 0.992) 30/32 <sup>n.s</sup> |
| **Induced sputum versus BAL** |  |  |
| **Induced sputum** | 1 (0.478, 1) 5/5 <sup>n.s</sup> | 1 (0.884, 1) 30/30 <sup>n.s</sup> |
| **BAL** | 0.600 (0.147, 0.947) 3/5 <sup>n.s</sup> | 0.938 (0.792, 0.992) 30/32 <sup>n.s</sup> |
| **Bronchial aspirate versus BAL** |  |  |
| **Bronchial aspirate** | 0.974 (0.865, 0.999) 38/39 *** | 0.999 (0.995, 1.000) 1142/1143 ** |
| **BAL** | 0.564 (0.396, 0.722) 22/39 *** | 0.985 (0.977, 0.991) 1142/1159 ** |
| All p.values in supplementary material |  |  |

These data suggest, no significant difference in the sensitivity of PCR between sputum and induced sputum when the patient can produce spontaneous sputum. In contrast, bronchial aspirate displays higher sensitivity than sputum and BAL. PCR displayed a high specificity for all the respiratory specimens.

### 3.6 Performance of culture according to the type of respiratory specimen

Using the classical “per specimen” approach, the sensitivity of culture when considering all respiratory specimen was 0.988 (0.978, 0.994). The sensitivities of culture for sputum, induced sputum, bronchial aspirate and BAL were 0.993 (0.981, 0.998), 0.980 (0.931, 0.998), 0.965 (0.919, 0.988), and 1 (0.961, 1).

Using the “per patient” comparison of paired specimens, no significant difference in sensitivity was seen between the different respiratory specimens expect a superiority of bronchial aspirate over BAL, 0.970 (0.914, 0.994) versus 0.636 (0.538, 0.731) (p. value < 0.0001) (Table 6,S6 and S11).

**Table 6.** Sensitivity of culture to predict tuberculosis according to the type of specimen using a 72-hours pairing window within the same patient.

|  |  |  |
| --- | --- | --- |
|  | Sensitivity | NPV |
| **Sputum versus induced sputum** |  |  |
| **Sputum** | 0.857 (0.697, 0.952) 30/35 <sup>n.s</sup> | 0.987 (0.969, 0.996) 368/373 <sup>n.s</sup> |
| **Induced sputum** | 0.800 (0.631, 0.916) 28/35 <sup>n.s</sup> | 0.981 (0.962, 0.992) 368/375 <sup>n.s</sup> |
| **Sputum versus bronchial aspirate** |  |  |
| **Sputum** | 0.613 (0.422, 0.782) 19/31 <sup>n.s</sup> | 0.964 (0.937, 0.981) 318/330 <sup>n.s</sup> |
| **Bronchial aspirate** | 0.903 (0.742, 0.980) 28/31 <sup>n.s</sup> | 0.991 (0.973, 0.998) 318/321 <sup>n.s</sup> |
| **Sputum versus BAL** |  |  |
| **Sputum** | 0.895 (0.669, 0.987) 17/19 <sup>n.s</sup> | 0.993 (0.974, 0.999) 268/270 <sup>n.s</sup> |
| **BAL** | 0.789 (0.544, 0.939) 15/19 <sup>n.s</sup> | 0.985 (0.963, 0.996) 268/272 <sup>n.s</sup> |
| **Induced sputum versus bronchial aspirate** |  |  |
| **Induced sputum** | 0.692 (0.386, 0.909) 9/13 <sup>n.s</sup> | 0.915 (0.796, 0.976) 43/47 <sup>n.s</sup> |
| **Bronchial aspirate** | 0.846 (0.546, 0.981) 11/13 <sup>n.s</sup> | 0.956 (0.849, 0.995) 43/45 <sup>n.s</sup> |
| **Induced sputum versus BAL** |  |  |
| **Induced sputum** | 0.769 (0.462, 0.950) 10/13 <sup>n.s</sup> | 0.929 (0.805, 0.985) 39/42 <sup>n.s</sup> |
| **BAL** | 0.462 (0.192, 0.749) 6/13 <sup>n.s</sup> | 0.848 (0.711, 0.937) 39/46 <sup>n.s</sup> |
| **Bronchial aspirate versus BAL** |  |  |
| **Bronchial aspirate** | 0.970 (0.914, 0.994) 96/99*** | 0.999 (0.997, 1.000) 3471/3474*** |
| **BAL** | 0.636 (0.538, 0.731) 63/99*** | 0.990 (0.986, 0.993) 3471/3507*** |
| All p.values details in supplementary material |  |  |

## 4. Discussion

The microbial diagnosis of tuberculosis is based on a combination of different microbiological tests that can be performed on different types of clinical samples. We aimed to identify the most efficient tests and specimen in order to guarantee an ideal sensitivity and specificity and to limit the use of unnecessary tests.

### Smear independent diagnostic of tuberculosis

Our results confirm a limited sensitivity of smear microscopy (0.523). The specificity (0.994) remains high, probably because of the extremely low prevalence (0.036) of positive microscopy. Our data suggest a higher sensitivity of microscopy on sputum over the three other samples. This is probably a bias because patients for whom the microbiological diagnosis is not made on spontaneous sputum and who therefore need induced sputum or bronchial aspiration and BAL are patients with paucibacillary infections as previously reported by Cadena et al [17]. Using a pairwise comparison method to avoid the patient effect, we do not see any significant difference in sensitivity of microscopy between the various clinical specimens. With a time to result lower than 30 minutes, microscopy remains virtually the fastest diagnostic test. However, its sensitivity and specificity is limited and it requires a lot of work by specialized personnel and the performance of this test may vary depending on the experience of the examiner [8,18]. In a region with a low prevalence of tuberculosis, the question arises of the usefulness of this test. In 2016, we introduced in our laboratory a smear-independent algorithm for the diagnostic of tuberculosis [8]. For all the suspicion of tuberculosis the microbial diagnosis is initiated by PCR; microscopy were not achieved anymore in emergency but grouped once per day. In case of suspected pulmonary tuberculosis, we initiated microbial diagnostic by the rapid molecular test Xpert MTB/RIF further replaced by Xpert MTB/RIF Ultra which is used both to detect *M. tuberculosis* DNA and to address patient infectiousness based on the semi-quantitative result; microscopic analysis was still performed after treatment start, in particular to guide contact tracing and des-isolation decisions [5,8,19]. Another study on the diagnosis of NTM infection also suggested a limited added-value of microscopy when 16S broad range PCR for the detection of NTM is available [18]. Our data suggested that microscopy might be useful only for patients with a high pre-test probability of NTM infections, such as immunocompromised patients or patients with clinical and radiological suspicion of having NTM lung disease. In February 2020, we and other diagnostic laboratory experienced an important staff limitation triggered by the SARS-CoV-2 pandemics. Indeed, during the first wave of the SARS-CoV-2 pandemic, the biomedical technicians were reassigned for the management of the SARS-CoV-2 RT-PCR tests. In this context, we had to rapidly identify all the unnecessary analysis among which was smear microscopy. As an immediate response, we therefore push forward, in February 2020, the smear independent algorithm for the diagnostic of mycobacterial infection. Since then, microscopy for the detection of acid-fast bacilli is achieved only on specific request from clinicians or for patients with a confirmed diagnostic of tuberculosis; indeed, microscopy can be useful for treatment follow-up because PCR can remain positive for a long period in patient’s respiratory specimens even when a treatment is well conducted and for contact tracing investigations. Microscopy in addition to pan-mycobacterial PCR can also be requested when there is a high pre-test probability of NTM infection [18].

### Towards less cultures?

PCR has improved the diagnostic of tuberculosis with a lower limit of detection than microscopy and an increased specificity for PCR targeting specific *M. tuberculosis* DNA sequences such as the IS6110. Culture, the oldest microbiological test for tuberculosis, remains the reference method with the lowest limit of detection [2,3]. The performance of mycobacterial culture, sensitivity (98.8%) and specificity (100%), was calculated using a composite gold standard including all the microbiological tests as well as clinical data. We reported few patients with positive PCR but negative culture. It will therefore be difficult to do without culture. Further optimization strategies could be implemented by selecting the most performant tests on the most efficient clinical samples regarding pulmonary tuberculosis. Thus, we could consider only a combination of (i) samples for PCR-based diagnosis to have short time to results coupled to (ii) a selection of samples to perform the culture warranted to obtain strains for testing susceptibility towards anti-mycobacterial agents and also to guarantee an optimal sensitivity (with delayed results).

When pulmonary tuberculosis is suspected, sputum is the usual specimen that is collected. Regarding microscopy, as indicated above there is no significant difference between the different types of clinical samples. On the other hand, with regard to PCR and culture which are much more sensitive and reliable tests we observed differences between the clinical specimens. When looking at sputum and induced sputum we do not see a significant difference in terms of sensitivity for culture. Several studies reported an increased sensitivity of induced sputum over sputum [20,21]. Using the classical approach or the pairwise comparison in the same patient, we did not observe a significant increase in sensitivity with induced sputum compared to spontaneous sputum. However, the pairwise comparisons suggest an increase in the yield of positivity when performing the two specimens. This conclusion should be confirmed by further studies. BAL and bronchial aspirate are generally coupled. The pairwise comparison demonstrates that in the case of tuberculosis the bronchial aspirate (97% of sensitivity) outcompete BAL (63.6% of sensitivity) suggesting a limited added benefit of the BAL for the diagnosis of tuberculosis. Bronchial aspirate and BAL are invasive, which make prospective studies hardly conceivable. Therefore, this retrospective study, giving access to 3’570 pairwise comparison including these specimen provide important data on their performance. Bronchoscopy is not only useful for tuberculosis diagnosis but also to investigate other infectious or non-infectious disease [22]. For instance, BAL is a very good specimen for the diagnostic of fungal infection such as *Pneumocystis jirovecii* infections or *Aspergillus fumigatus* infection [23-25]. In view of these results, it is important to consider bronchial aspiration for the diagnosis of tuberculosis, confirming previous studies [26]. A first step would therefore be to always add a search for mycobacteria on the bronchial aspiration when it is missing in laboratory order. This is what we systematically do in our lab when it is missing. This study provides data for diagnostic stewardship and guidance for physicians and clinical microbiology laboratories. Such data could also help at defining diagnostic strategies in the setting of staff or reagants shortage or to reduce costs. Indeed, even in low prevalence and high-income country, the infrastructure and trained personnel for the diagnostic of tuberculosis is limited [27]. The SARS-CoV-2 pandemics negatively impacted tuberculosis control because of the mobilization of trained staff for other activities or because of the disruption of the supply chain of reagents and compounds for tuberculosis diagnostic and treatments [28,29]. In a context of shortage of reagants or other material for mycobacterial culture like the one we encountered during the COVID-19 pandemic, if a prioritization had to be made, it should be done for the benefit of bronchial aspiration; but this should be decided together with the clinician that can help guiding the decision by providing clinical information for each case.

### Limits of the study and perspectives

Molecular diagnostic significantly improved the microbial diagnostic of tuberculosis, in particular the initial diagnostic, but is not yet generalized worldwide [30], mainly for economical reasons. To assess the real economic impact of the management of tuberculosis a cost-benefit analysis for the full replacement of microscopy in favor of PCR should be performed since this study demonstrates the effectiveness of PCR over microscopy [31]. Such analysis should incorporate that, in hospital setting, patients might be isolated in specific negative pressure chambers for the duration of the investigation. Such cost-benefit study should also address the risk of nosocomial infection due to delayed diagnostic. Future studies should account for the evolution in practices that may have occurred over the ten years (2008-2018) of the study. It would be worthwhile to relate the data with the evolution of protocols and guidelines that were introduced during the studied period in order not to only add diagnostic tools but also to stop the not useful approaches. This study will also permit to address the dependence between the tests results and many other parameters such as the number of tests, the quality of the clinical specimens and patients characteristics. This will be particularly useful for results interpretation, in particular negative results. Finally, this study is based on a large amount of data over a long period, which was made possible by the fact that all the microbial result are in our LIS since 1995. Although, it may not be the case even in high income country labs, comparison of these results with those obtained at other medical centers should be performed with the view of cross-validating the robustness of the present results.

## Conclusions

This study demonstrates that many improvements have been made in the microbiological diagnosis of tuberculosis. There is no doubt about the added value of molecular diagnosis compared to microscopy to initiate the diagnosis of tuberculosis. The limit of a generalization of independent algorithms in microscopy lies in the access to molecular diagnosis. New technologies such as the GeneXpert, which are supposed to solve this problem, are not yet generalized. Regarding tuberculosis, the limit in this case is not technological but again, economical. This study provides data for diagnostic stewardship and for editing guidelines and diagnosis protocols with the purpose to reduce the medical, social and economic costs associated with tuberculosis. Indeed, even in low prevalence and high-income country, the infrastructure and trained personnel for the diagnostic of tuberculosis is limited. Therefore, there is a need to identify the most efficient tests and specimens in order to guarantee the sensitivity and specificity and to limit the number of unnecessary tests. In addition, it provides analytical strategies that may also be suitable for the study of other infectious diseases.

## Data Availability

Data produced in the present study were obtained within the frame of an ethical autorisation; they would be available upon reasonable request to the authors and within the frame of another ethical autorisation only

## 5. Acknowledgements

We are thankful to the Laboratory of Molecular Diagnostic and Mycobacteria of the Institute of Microbiology of the University of Lausanne and Lausanne University Hospital that perform the conventional and molecular analysis for the diagnosis of tuberculosis.

## 8. Supplementary material

**Table S1.**
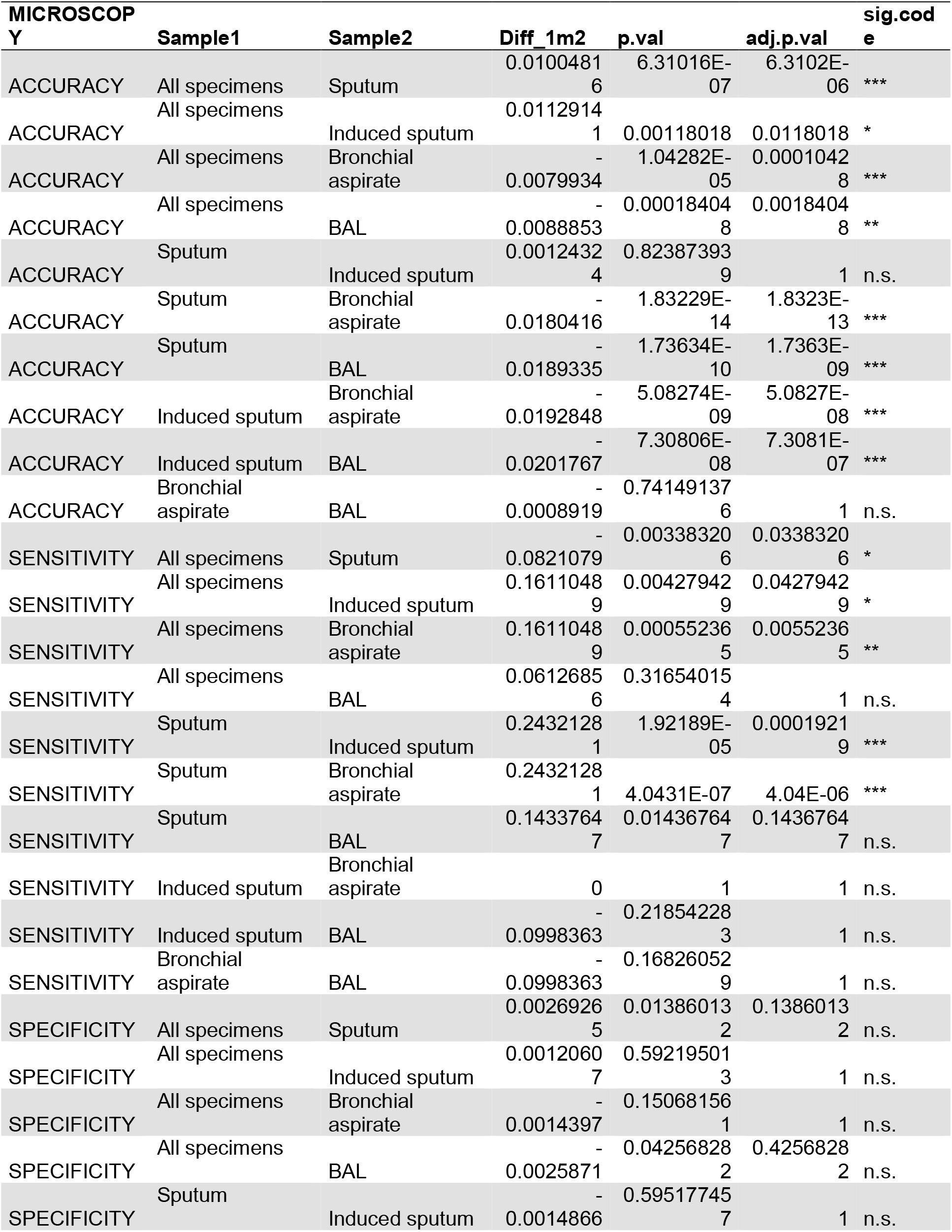
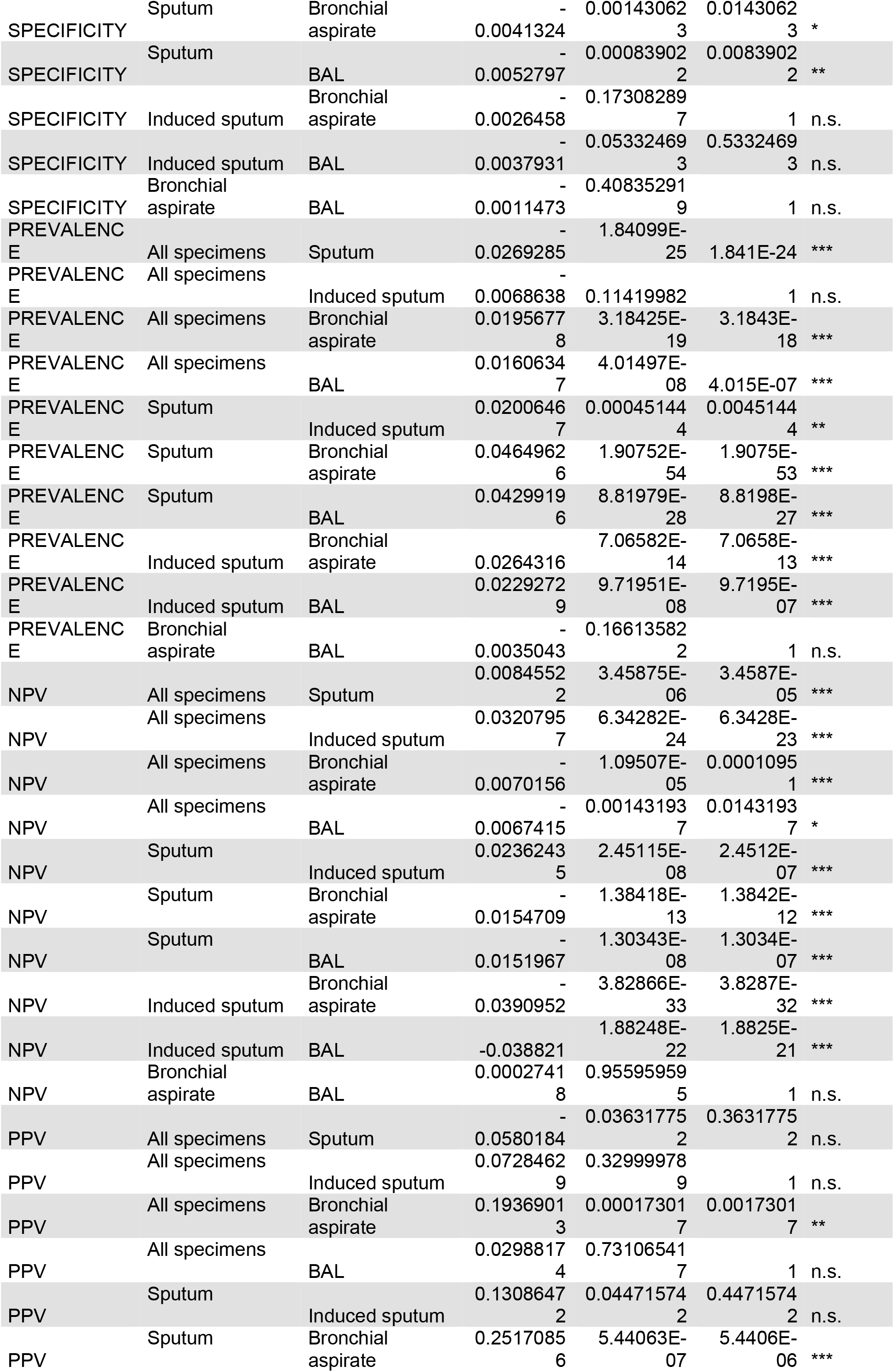
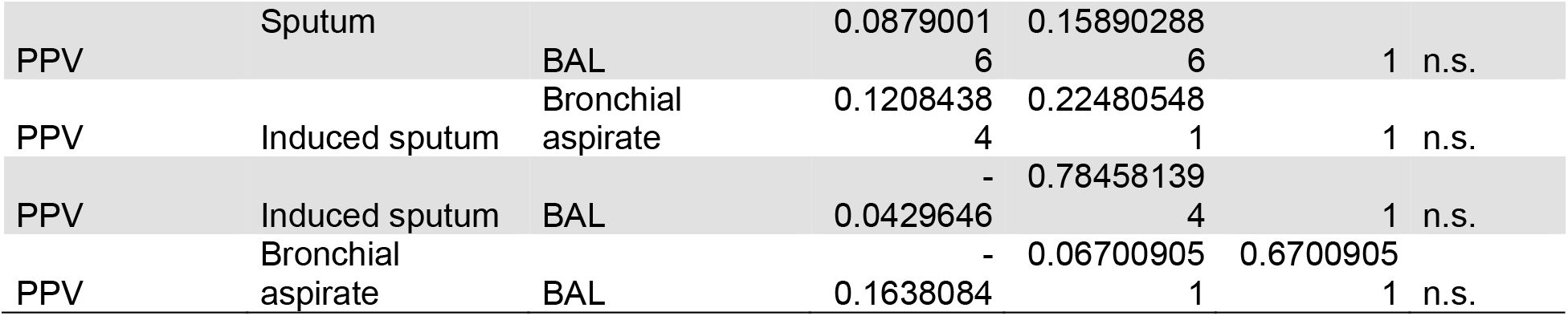
P-values for Table 1, performance of smear microscopy according of the type of specimen.

**Table S2.** P-values for Table 2, performance of PCR according of the type of specimen.

| PCR | Sample1 | Sample2 | Diff_1m2 | p.val | adj.p.val | sig.code |
| --- | --- | --- | --- | --- | --- | --- |
| ACCURACY | All specimens | Sputum | 0.00287789 | 0.304418176 | 1 | n.s. |
| ACCURACY | All specimens | Induced sputum | 0.01147752 | 0.019512972 | 0.19512972 | n.s. |
| ACCURACY | All specimens | Bronchial aspirate | -0.0040849 | 0.123765813 | 1 | n.s. |
| ACCURACY | All specimens | BAL | -0.0034643 | 0.283404464 | 1 | n.s. |
| ACCURACY | Sputum | Induced sputum | 0.00859963 | 0.167946429 | 1 | n.s. |
| ACCURACY | Sputum | Bronchial aspirate | -0.0069627 | 0.030595877 | 0.30595877 | n.s. |
| ACCURACY | Sputum | BAL | -0.0063422 | 0.092258909 | 0.92258909 | n.s. |
| ACCURACY | Induced sputum | Bronchial aspirate | -0.0155624 | 0.001625285 | 0.01625285 | * |
| ACCURACY | Induced sputum | BAL | -0.0149418 | 0.007047431 | 0.07047431 | n.s. |
| ACCURACY | Bronchial aspirate | BAL | 0.00062059 | 0.975120507 | 1 | n.s. |
| SENSITIVITY | All specimens | Sputum | -0.0232511 | 0.562771431 | 1 | n.s. |
| SENSITIVITY | All specimens | Induced sputum | 0.15514784 | 0.033580028 | 0.33580028 | n.s. |
| SENSITIVITY | All specimens | Bronchial aspirate | -0.0387297 | 0.467749275 | 1 | n.s. |
| SENSITIVITY | All specimens | BAL | 0.03874573 | 0.630382277 | 1 | n.s. |
| SENSITIVITY | Sputum | Induced sputum | 0.1783989 | 0.017069317 | 0.17069317 | n.s. |
| SENSITIVITY | Sputum | Bronchial aspirate | -0.0154787 | 0.863947971 | 1 | n.s. |
| SENSITIVITY | Sputum | BAL | 0.06199678 | 0.402691103 | 1 | n.s. |
| SENSITIVITY | Induced sputum | Bronchial aspirate | -0.1938776 | 0.020902399 | 0.20902399 | n.s. |
| SENSITIVITY | Induced sputum | BAL | -0.1164021 | 0.308470102 | 1 | n.s. |
| SENSITIVITY | Bronchial aspirate | BAL | 0.07747543 | 0.343242299 | 1 | n.s. |
| SPECIFICITY | All specimens | Sputum | -0.0001495 | 1 | 1 | n.s. |
| SPECIFICITY | All specimens | Induced sputum | -0.0001495 | 1 | 1 | n.s. |
| SPECIFICITY | All specimens | Bronchial aspirate | -0.0001495 | 1 | 1 | n.s. |
| SPECIFICITY | All specimens | BAL | -0.0001495 | 1 | 1 | n.s. |
| SPECIFICITY | Sputum | Induced sputum | 0 | NA | NA |  |
| SPECIFICITY | Sputum | Bronchial aspirate | 0 | NA | NA |  |
| SPECIFICITY | Sputum | BAL | 0 | NA | NA |  |
| SPECIFICITY | Induced sputum | Bronchial aspirate | 0 | NA | NA |  |
| SPECIFICITY | Induced sputum | BAL | 0 | NA | NA |  |
| SPECIFICITY | Bronchial aspirate | BAL | 0 | NA | NA |  |
| PREVALENC E | All specimens | Sputum | -0.0242457 | 1.85046E-05 | 0.00018505 | *** |
| PREVALENC E | All specimens | Induced sputum | -0.012415 | 0.24121711 | 1 | n.s. |
| PREVALENC E | All specimens | Bronchial aspirate | 0.01343573 | 0.015178942 | 0.15178942 | n.s. |
| PREVALENC E | All specimens | BAL | 0.0229057 | 0.000252936 | 0.00252936 | ** |
| PREVALENC E | Sputum | Induced sputum | 0.01183074 | 0.372502519 | 1 | n.s. |
| PREVALENC E | Sputum | Bronchial aspirate | 0.03768143 | 1.03816E-07 | 1.0382E-06 | *** |
| PREVALENC E | Sputum | BAL | 0.04715139 | 1.47378E-09 | 1.4738E-08 | *** |
| PREVALENC E | Induced sputum | Bronchial aspirate | 0.02585069 | 0.011433057 | 0.11433057 | n.s. |
| PREVALENC E | Induced sputum | BAL | 0.03532066 | 0.000420219 | 0.00420219 | ** |
| PREVALENC E | Bronchial aspirate | BAL | 0.00946997 | 0.157733443 | 1 | n.s. |
| NPV | All specimens | Sputum | 0.0355276 | 1.72723E-11 | 1.7272E-10 | *** |
| NPV | All specimens | Induced sputum | -0.0212302 | 0.016901135 | 0.16901135 | n.s. |
| NPV | All specimens | Bronchial aspirate | -0.0448089 | 3.59424E-23 | 3.5942E-22 | *** |
| NPV | All specimens | BAL | -0.0369566 | 1.0787E-11 | 1.0787E-10 | *** |
| NPV | Sputum | Induced sputum | -0.0567578 | 9.29799E-07 | 9.298E-06 | *** |
| NPV | Sputum | Bronchial aspirate | -0.0803365 | 8.28082E-41 | 8.2808E-40 | *** |
| NPV | Sputum | BAL | -0.0724842 | 1.81678E-23 | 1.8168E-22 | *** |
| NPV | Induced sputum | Bronchial aspirate | -0.0235787 | 4.52576E-11 | 4.5258E-10 | *** |
| NPV | Induced sputum | BAL | -0.0157264 | 0.006062577 | 0.06062577 | n.s. |
| NPV | Bronchial aspirate | BAL | 0.00785225 | 0.00031041 | 0.0031041 | ** |
| PPV | All specimens | Sputum | -0.0031153 | 1 | 1 | n.s. |
| PPV | All specimens | Induced sputum | -0.0031153 | 1 | 1 | n.s. |
| PPV | All specimens | Bronchial aspirate | -0.0031153 | 1 | 1 | n.s. |
| PPV | All specimens | BAL | -0.0031153 | 1 | 1 | n.s. |
| PPV | Sputum | Induced sputum | 0 | NA | NA |  |
| PPV | Sputum | Bronchial aspirate | 0 | NA | NA |  |
| PPV | Sputum | BAL | 0 | NA | NA |  |
| PPV | Induced sputum | Bronchial aspirate | 0 | NA | NA |  |
| PPV | Induced sputum | BAL | 0 | NA | NA |  |
| PPV | Bronchial aspirate | BAL | 0 | NA | NA |  |

**Table S3.**
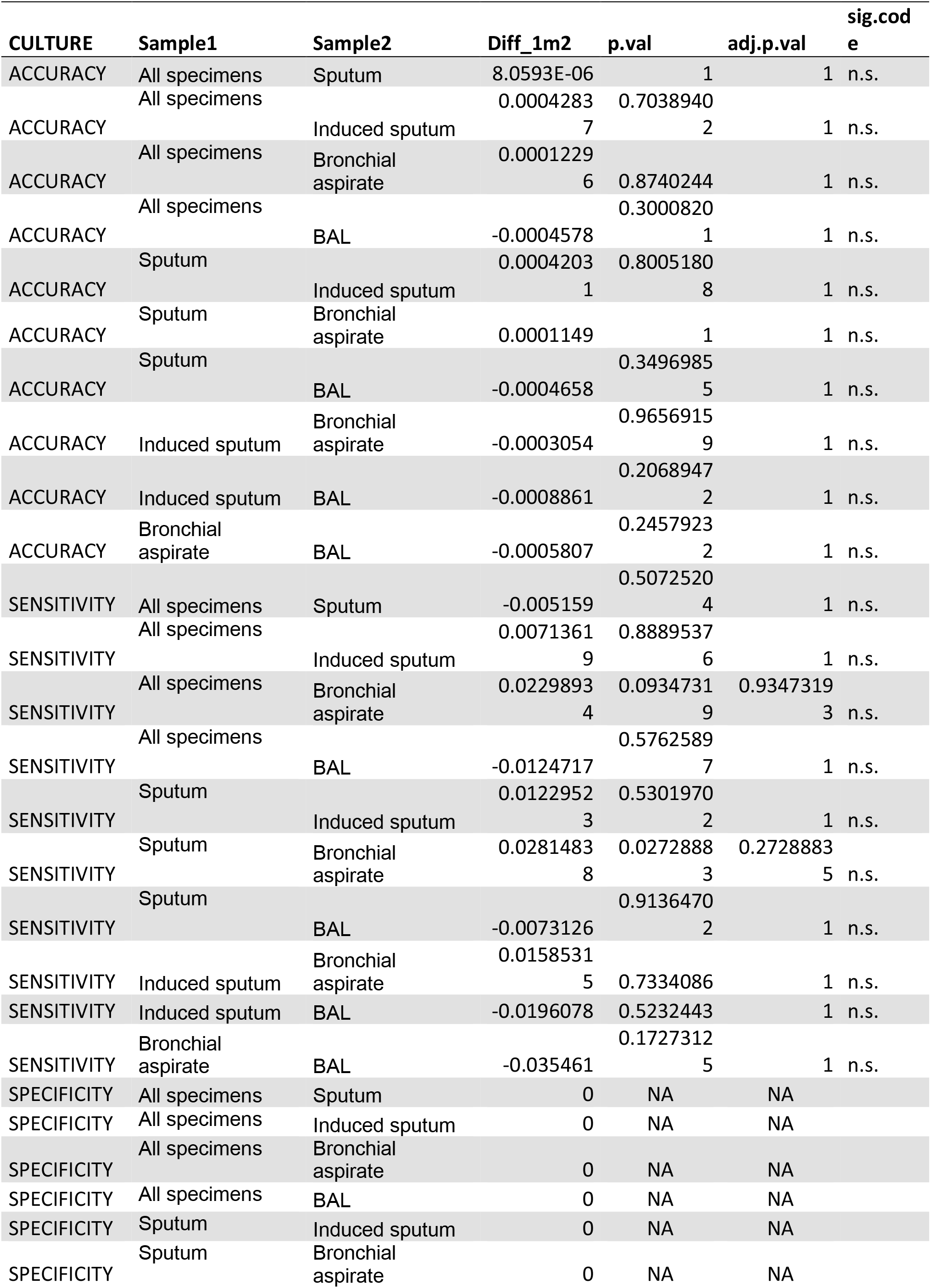
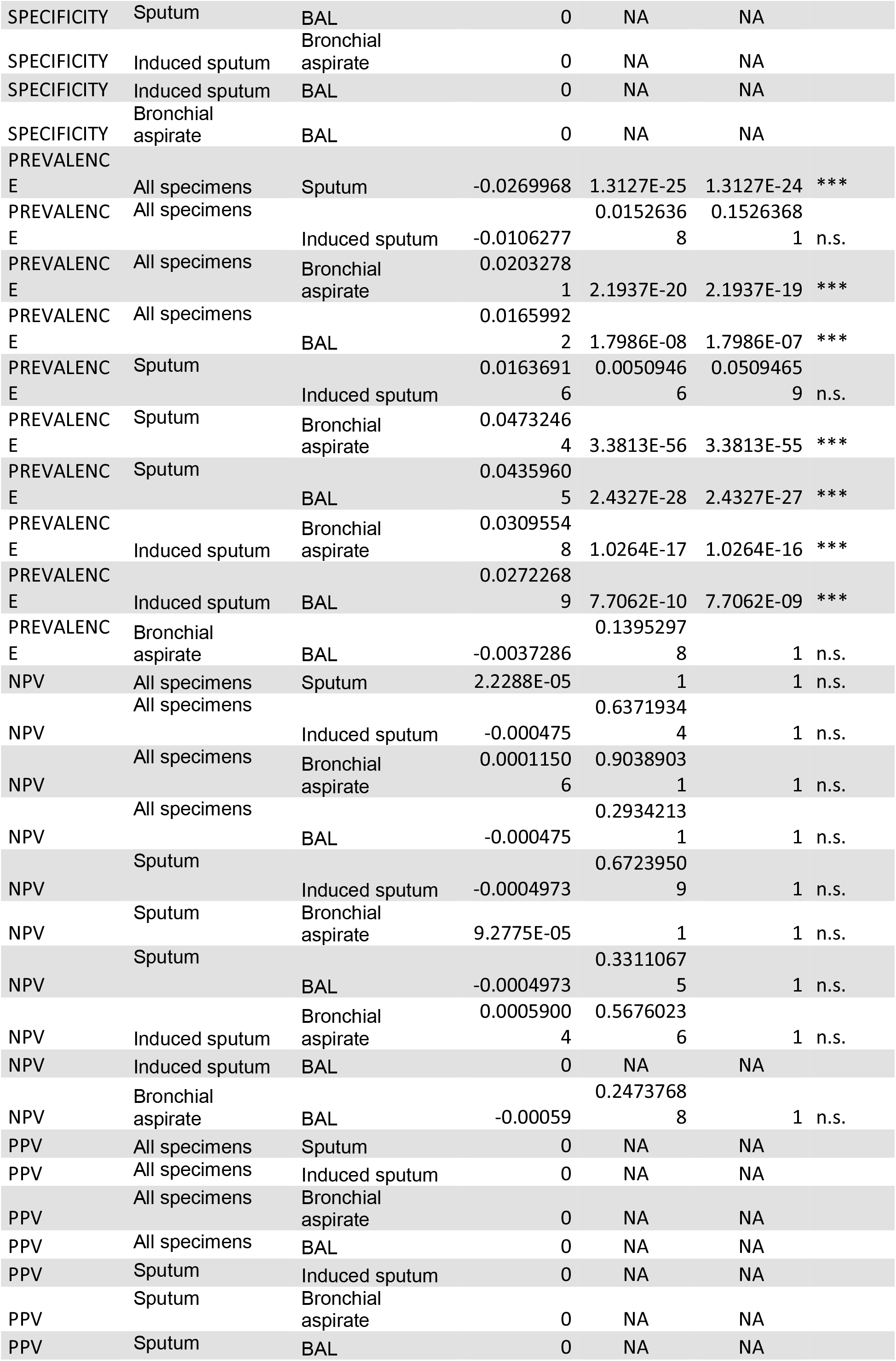
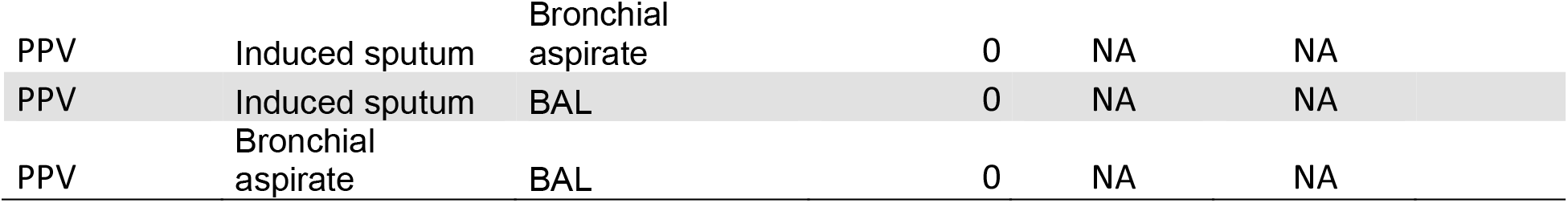
P-values for Table 3, performance of culture according of the type of specimen.

**Table S4.** P-values for Table 4, performance of smear microscopy according of the type of specimen.

|  |  |  |  |  |  |  |
| --- | --- | --- | --- | --- | --- | --- |
| Pairwise Comparison Direct |  |  |  |  |  |  |
| Exam | Sample1 | Sample2 | Diff_1m2 | p.val | adj.p.val | sig.code |
| SENSITIVITY | Sputum | Induced sputum | 0.15384615 | 0.58674615 | 1 | n.s. |
| SENSITIVITY | Sputum | Bronchial aspirate | -0.1538462 | 0.64159157 | 1 | n.s. |
| SENSITIVITY | Sputum | BAL | 0.25 | 0.4496918 | 1 | n.s. |
| SENSITIVITY | Induced sputum | Bronchial aspirate | 0 | 1 | 1 | n.s. |
| SENSITIVITY | Induced sputum | BAL | -0.3333333 | 1 | 1 | n.s. |
| SENSITIVITY | Bronchial aspirate | BAL | 0.16666667 | 0.10322732 | 0.61936393 | n.s. |
| NPV | Sputum | Induced sputum | 0.00507604 | 0.61977665 | 1 | n.s. |
| NPV | Sputum | Bronchial aspirate | -0.0058305 | 0.68707568 | 1 | n.s. |
| NPV | Sputum | BAL | 0.0070922 | 0.48184948 | 1 | n.s. |
| NPV | Induced sputum | Bronchial aspirate | 0 | 1 | 1 | n.s. |
| NPV | Induced sputum | BAL | -0.0188537 | 1 | 1 | n.s. |
| NPV | Bronchial aspirate | BAL | 0.0019746 | 0.14625681 | 0.87754084 | n.s. |

**Table S5.**
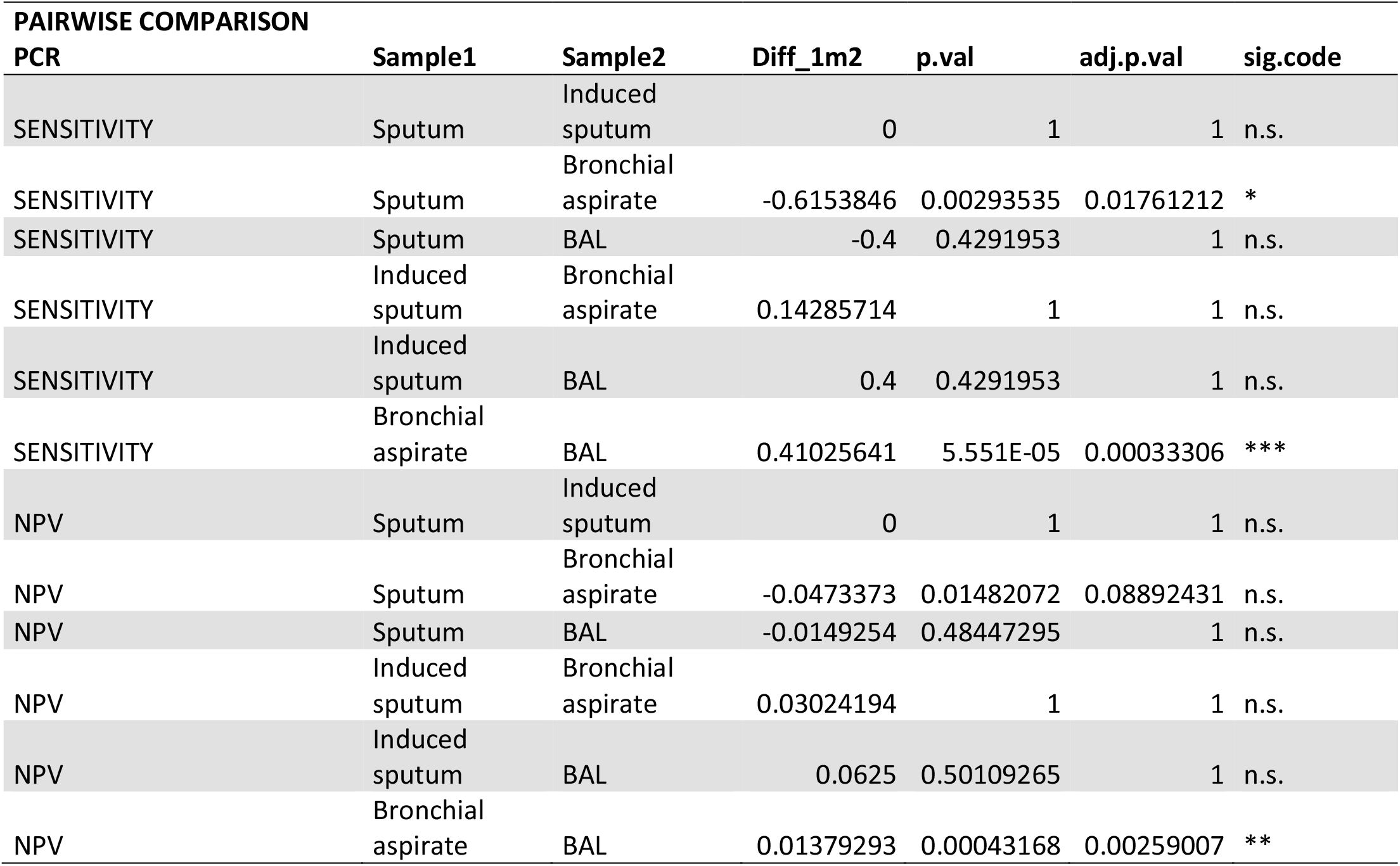
P-values for Table 5, sensitivity of PCR to predict tuberculosis according to the type of specimen using a 72 72-hours pairing window in the same patient.

| Table S5. P.values for table 5, sensitivity of PCR to predict tuberculosis according to the type of specimen using a 72-hours pairing window in the same patient.. |  |  |  |  |  |  |
| --- | --- | --- | --- | --- | --- | --- |
| PAIRWISE COMPARISON |  |  |  |  |  |  |
| PCR | Sample1 | Sample2 | Diff_1m2 | p.val | adj.p.val | sig.code |
| SENSITIVITY | Sputum | Induced sputum | 0 | 1 | 1 | n.s. |
| SENSITIVITY | Sputum | Bronchial aspirate | -0.6153846 | 0.00293535 | 0.01761212 | * |
| SENSITIVITY | Sputum | BAL | -0.4 | 0.4291953 | 1 | n.s. |
| SENSITIVITY | Induced sputum | Bronchial aspirate | 0.14285714 | 1 | 1 | n.s. |
| SENSITIVITY | Induced sputum | BAL | 0.4 | 0.4291953 | 1 | n.s. |
| SENSITIVITY | Bronchial aspirate | BAL | 0.41025641 | 5.551E-05 | 0.00033306 | *** |
| NPV | Sputum | Induced sputum | 0 | 1 | 1 | n.s. |
| NPV | Sputum | Bronchial aspirate | -0.0473373 | 0.01482072 | 0.08892431 | n.s. |
| NPV | Sputum | BAL | -0.0149254 | 0.48447295 | 1 | n.s. |
| NPV | Induced sputum | Bronchial aspirate | 0.03024194 | 1 | 1 | n.s. |
| NPV | Induced sputum | BAL | 0.0625 | 0.50109265 | 1 | n.s. |
| NPV | Bronchial aspirate | BAL | 0.01379293 | 0.00043168 | 0.00259007 | ** |

**Table S6.**
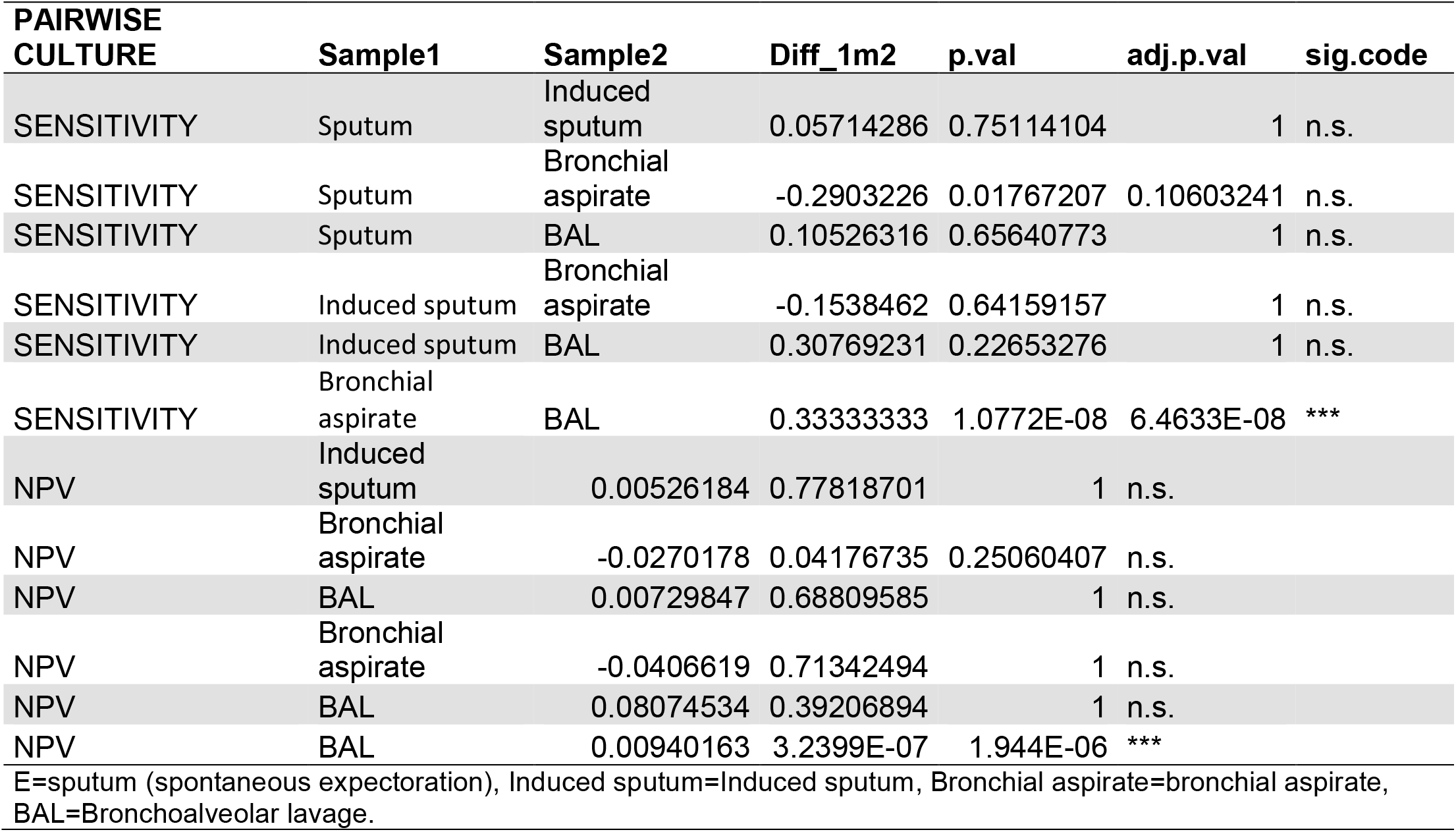
P.values for table 1, sensitivity of culture to predict tuberculosis according to the type of specimen using a 72-hour pairing window within the same patient.

**Table S7.** Performance of in-house Taqman PCR and Xpert for the diagnostic of pulmonary tuberculosis.

|  | Accuracy | Sensitivity | Specificity | Prevalence | PPV | NPV |
| --- | --- | --- | --- | --- | --- | --- |
| **In-house TaqMan PCR** | 0.991<br>(0.988, 0.994)<br>5493/5542 | 0.799<br>(0.743, 0.848)<br>191/239 | >0.999<br>(0.999, 1)<br>5302/5303 | 0.043<br>(0.038, 0.049)<br>239/5542 | 0.995<br>(0.971, 1)<br>191/192 | 0.991<br>(0.988, 0.993)<br>5302/5350 |
| **Xpert** | 0.975<br>(0.967, 0.981)<br>1917/1967 | 0.812<br>(0.760, 0.858)<br>212/261 | 0.999<br>(0.997, 1)<br>1705/1706 | 0.133<br>(0.118, 0.148)<br>261/1967 | 0.995<br>(0.974, 1)<br>212/213 | 0.972<br>(0.963, 0.979)<br>1705/1754 |

**Table S8.** Kappa with 95% confidence intervals.

|  | Microscopy | PCR | Culture |
| --- | --- | --- | --- |
| Sputum vs Induced sputum | 0.73 (0.54,0.92) | 0.49 (-0.11,1) | 0.78 (0.66,0.9) |
| Sputum vs Bronchial aspirate | 0.79 (0.62,0.97) | 0.54 (0.26,0.81) | 0.66 (0.50,0.82) |
| Sputum vs BAL | 0.93 (0.80,1) | 0.74 (0.40,1) | 0.80 (0.65,0.96) |
| Induced sputum vs Bronchial aspirate | -0.02 (-0.04,0.01) | 0.68 (0.34,1) | 0.64 (0.37,0.90) |
| Induced sputum vs BAL | -0.02 (-0.05,0.01) | 0.72 (0.36,1) | 0.27 (-0.06,0.60) |
| Bronchial aspirate vs BAL | 0.72 (0.62,0.82) | 0.69 (0.56,1) | 0.75 (0.67,0.83) |

**Table S9.**
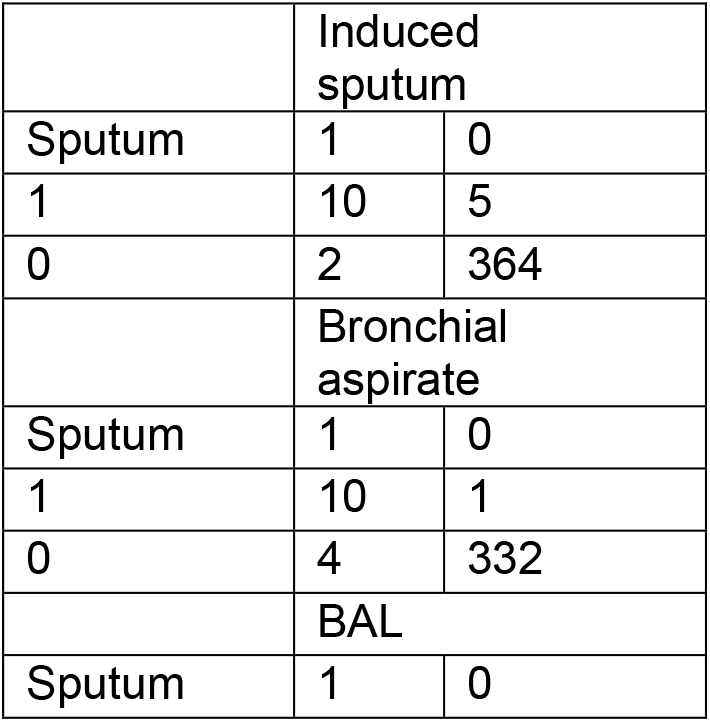
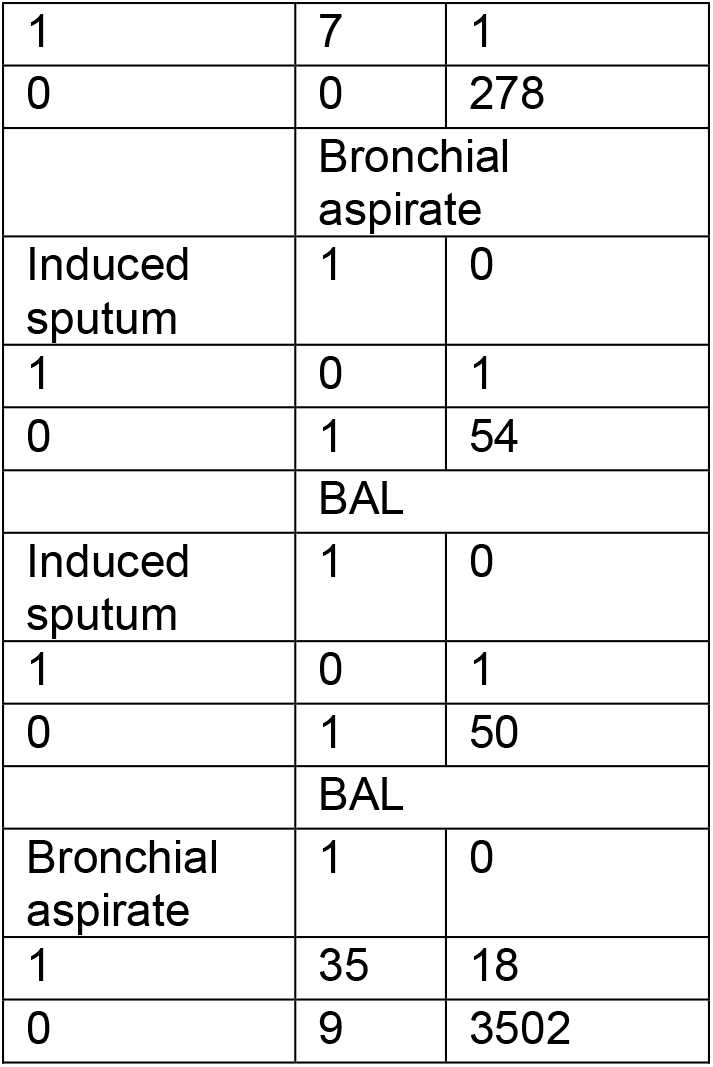
Contingency table for kappa: microscopy.

**Table S10.**
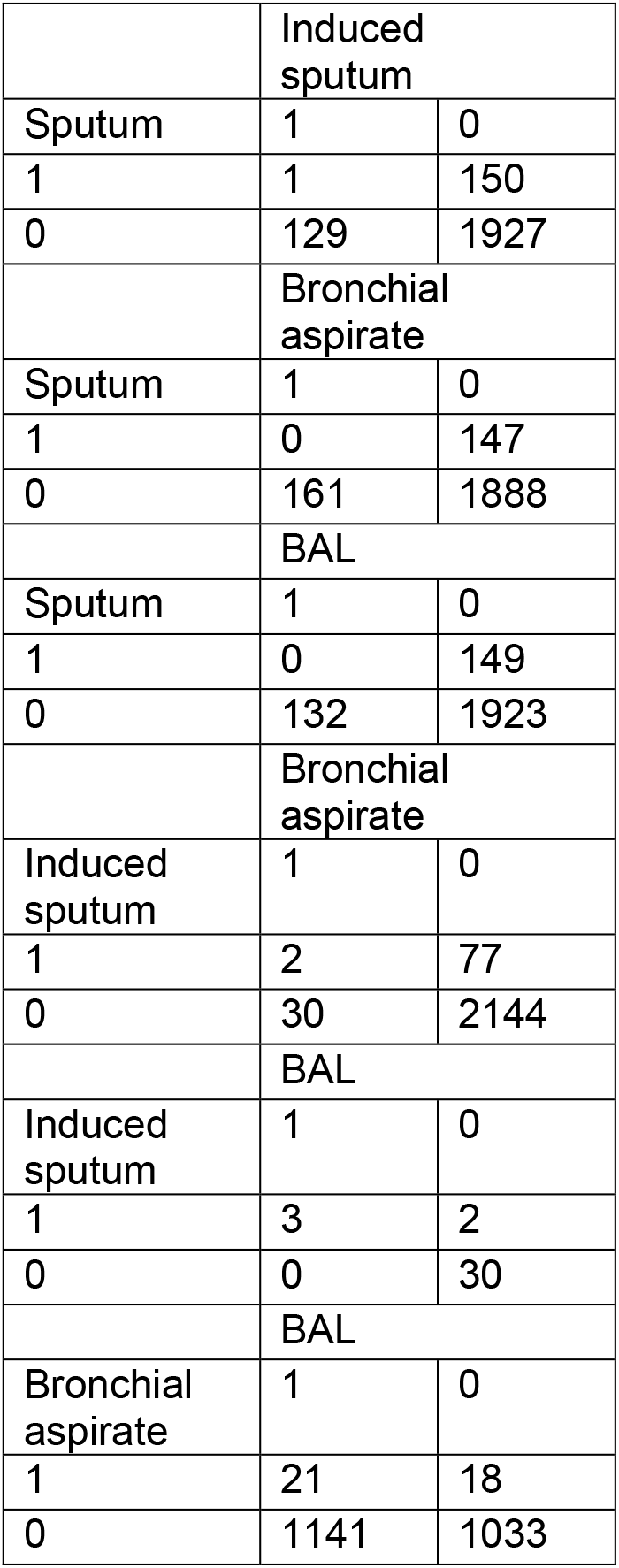
Contingency table for kappa: PCR.

**Table S11.**
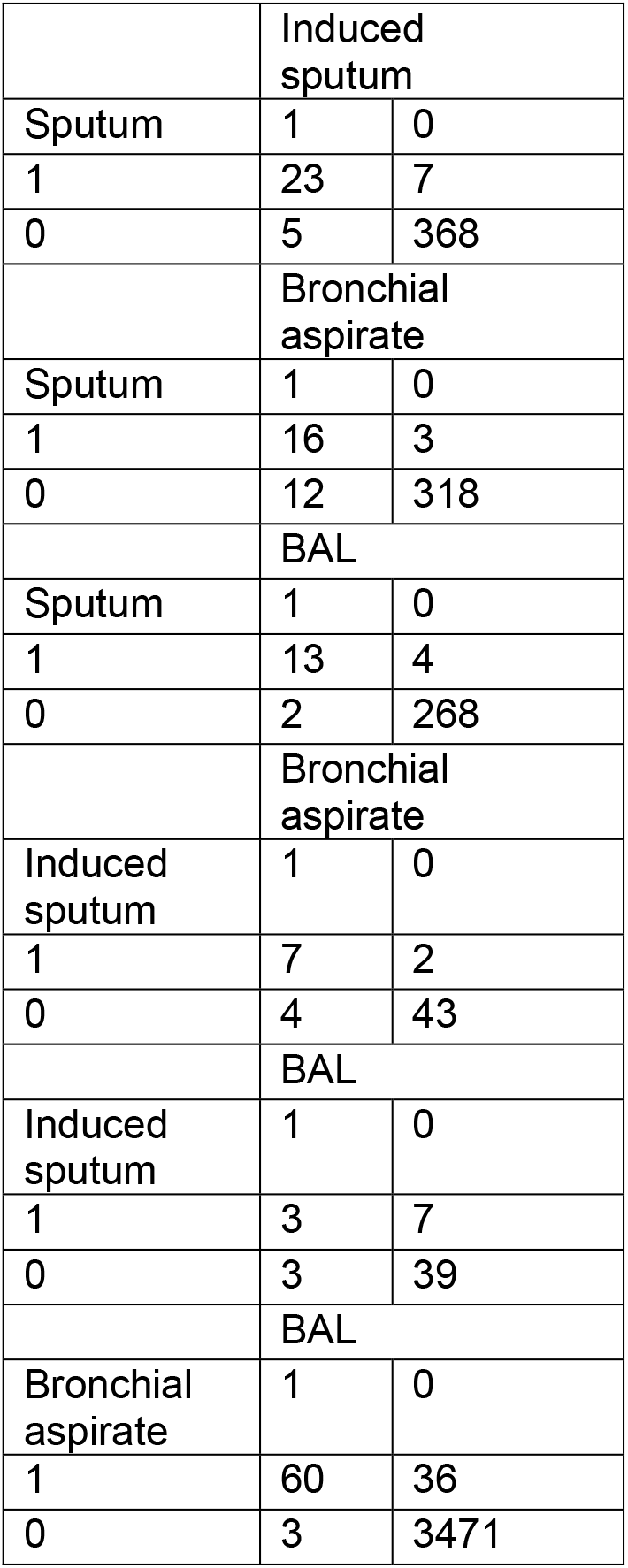
Contingency table for kappa: culture.

|  |  |  |
| --- | --- | --- |
|  | Induced sputum |  |
| Sputum | 1 | 0 |
| 1 | 23 | 7 |
| 0 | 5 | 368 |
|  | Bronchial aspirate |  |
| Sputum | 1 | 0 |
| 1 | 16 | 3 |
| 0 | 12 | 318 |
|  | BAL |  |
| Sputum | 1 | 0 |
| 1 | 13 | 4 |
| 0 | 2 | 268 |
|  | Bronchial aspirate |  |
| Induced sputum | 1 | 0 |
| 1 | 7 | 2 |
| 0 | 4 | 43 |
|  | BAL |  |
| Induced sputum | 1 | 0 |
| 1 | 3 | 7 |
| 0 | 3 | 39 |
|  | BAL |  |
| Bronchial aspirate | 1 | 0 |
| 1 | 60 | 36 |
| 0 | 3 | 3471 |

## Notes

### Competing Interest Statement

The authors have declared no competing interest.

### Author Declarations

This study was approved by the relevant ethics committee, the cantonal Ethics Committee on human research of the Vaud Canton, Switzerland "Commission Cantonale d Ethique de la Recherche sur l Etre Humain du Canton de Vaud, Suisse, cer-vd.ch" (Approval number CER-2020-00136)

### Summary of Updates

The word "speminen" was missing in the title that now reads "Performance of microbiological tests for tuberculosis diagnostic according to the type of respiratory specimen: a 10-year retrospective study".

